# Elevated heart rate after non-cardiac surgery: post-hoc analysis of a prospective observational cohort study

**DOI:** 10.1101/19009530

**Authors:** Amour B. U. Patel, Shaun M. May, Anna Reyes, Gladys Martir, David Brealey, Robert C. M. Stephens, Tom E. F. Abbott, Gareth L. Ackland

## Abstract

**Background:** Elevated heart rate (HR) is associated with accelerated mortality and independently predicts poorer outcomes in patients discharged from hospital after myocardial infarction and/or heart failure. We examined whether resting HR measured within 24 hours of hospital discharge following elective non-cardiac surgery was elevated compared to preoperative values. We also investigated the relationship between changes in HR with and/or autonomic function associated with morbidity after surgery.

**Methods:** We conducted a post-hoc analysis of HR data obtained in a prospective observational cohort study of patients ≥18years in whom serial Holter-based measurements of cardiac autonomic activity were made before, and for 48h after, surgery. The primary outcome was absolute discharge HR (beats minute^-1^), recorded at rest before hospital discharge. We examined the association between quartiles of discharge HR and autonomic measures (time/frequency domain heart rate variability) associated with morbidity (defined by Postoperative morbidity survey).

**Results:** In 157 patients (66 (42%) male; age 67(9) years), HR at hospital discharge (range: 53-122) increased by 5 beats minute^-1^ (95%CI:3–7;p<0.001) compared to preoperative values. Patients in the upper quartile of discharge HR (≥85bpm) were more likely to sustain pulmonary (odds ratio (OR):2.18 (95%CI:1.07-4.44);p=0.03) and infectious (OR:2.31 (95%CI:1.13-4.75);p=0.02) morbidity within seven days of surgery, compared to lower quartiles. Pulmonary/infectious morbidity was associated with loss of cardiac vagal activity.

**Conclusions:** Heart rate on discharge from hospital following major elective non-cardiac surgery is frequently elevated and is promoted by morbidity associated with reductions in cardiac vagal activity.

## Introduction

In mammals, there is an inverse semilogarithmic relation between heart rate and life expectancy suggesting that for each species, there is a predetermined number of heart beats in a lifetime (1). Elevated heart rate is a consistent, robust predictor of morbidity and mortality, including in otherwise healthy individuals (2). Several prospective studies have identified higher heart rate at discharge from hospital as an independent prognostic predictor of morbidity and mortality in patients treated for acute myocardial infarction (3,4,5), and heart failure (2,6). For every 10 beats minute^-1^ (bpm) increase in heart rate, mortality increases by 16% in patients with heart failure (7). Before non-cardiac surgery, resting heart rate >87 beats minute^-1^ is associated with myocardial injury and poorer outcome (8,9).

A prototypical feature of the surgical stress response is loss of cardiac vagal activity, leading to higher resting heart rate (10). However, it is unknown whether increases in perioperative heart rate persist beyond the first 72h after non-cardiac surgery. If so, heart rate may be an overlooked modifiable risk factor for morbidity and/or mortality after discharge from hospital following surgery. Here, we hypothesised that elevation in resting heart rate after surgery persists until at least discharge from hospital. We performed a post-hoc analysis of a single-centre observational cohort study of non-cardiac surgical patients to determine whether cardiac vagal function and/or morbidity after surgery were associated with higher heart rate at the time of discharge from hospital.

## Methods

### Study design

This was a post-hoc analysis of a single-centre, prospective, observational mechanistic cohort study conducted at University College London Hospital. The study protocol was approved by a research ethics committee (MREC:16/LO/06/35) and conformed to the principles of the Helsinki declaration and the Research Governance Framework. Written informed consent was obtained from all participants for the collection of routinely collected clinical data prior to surgery. The Strengthening and Reporting of Observational Studies in Epidemiology (STROBE) guidelines (11) (STROBE statement in Supplementary Material) were followed.

### Inclusion and exclusion criteria

Patients aged ≥18 years undergoing elective non-cardiac surgery under general anaesthesia, regional anaesthesia, or both with a planned overnight stay in hospital were eligible for recruitment, provided they satisfied the following criteria: American Society Anaesthesiology (ASA) score ≥3, major surgery with expected surgical operating time ≥ 120 minutes. Patients with atrial and/or ventricular arrhythmias, (including permanent or temporary pacemaker in-situ), on discharge or day of surgery, or who refused consent, were excluded.

### Heart rate measurement

Resting preoperative heart rate values were measured once in the sitting position as part of the patients’ routine pre-assessment clinic visit (Carescape V100 Dinamap Vital Signs Monitor, GE Healthcare, UK). On the morning of surgery, continuous three lead electrocardiogram (ECG) recordings were made in a quiet temperature-controlled room. Resting preoperative heart rate values were measured again as the mean heart rate during this recording. These measurements were repeated at 24 hours and 48 hours after surgery at the same time of day. Resting discharge heart rate was measured within 24h of discharge in the sitting position by nursing staff (Carescape V100 Dinamap Vital Signs Monitor, GE Healthcare, UK) and recorded on National Early Warning Score observation charts. Three lead electrocardiogram (ECG) recordings were made at a sampling rate of 1024Hz (Lifecard CF digital Holter monitors, Spacelabs Healthcare, Hertford, UK).

### Assessment of cardiac autonomic activity-time and frequency domain measures

R-R intervals from ECG data were captured using Kubios heart rate variability (HRV) Premium software Version 3.1.0 (Kubios, Kuopio, Finland), as described previously (12). Holter data cleaning and analyses were performed blinded to perioperative data and outcomes. HRV analysis was performed for five-minute segments according to Taskforce guidelines (13). Serial measurements of cardiac autonomic activity were quantified by well-established measures of autonomic control of heart rate; time domain, frequency domain and orthostatic modulation of heart rate (further detailed methodology in Supplementary Material). We used the square root of the mean of the sum of the squares of the successive differences between adjacent beat-to beat intervals (RMSSD) and Standard Deviation of the N-N interval (SDNN) as time domain measures. We also analysed frequency domain measures, using high frequency (0.15-0.4Hz) power spectral analysis of R-R interval time series as a measure of cardiac parasympathetic activity. Low frequency (0.04-0.15Hz) power spectral analysis was also assessed as a measure of arterial baroreflex activity. Log transformation of these measures was performed.

### Perioperative management

Consultant doctors performed surgery and anaesthesia, and intraoperative management was of the usual standard of care. Postoperative care was performed as per the local enhanced recovery programme guidelines. All clinicians were blinded to Holter data and outcomes.

### Baseline characteristics

Baseline data were obtained through chart abstraction and standardized interviews by trained hospital research staff at the first preoperative encounter, discharge and most recent postoperative encounter. The baseline characteristics recorded are detailed in the CRF (Supplementary data):

### Postoperative morbidity

Postoperative morbidity was recorded using the Postoperative Morbidity Survey (POMS), which is an 18-item survey that investigates nine domains of postoperative morbidity (Supplementary Table 1) (14).

### Exposure of interest

The exposure of interest was the absolute heart rate value. Resting preoperative heart rate was defined as the heart rate value taken during the preoperative assessment clinic visit.

### Primary outcome

The primary outcome was the absolute resting heart rate within 24 hours of discharge from hospital.

### Secondary analyses

We also quantified the absolute change in resting heart rate from preoperative to within 24 hours of discharge for each patient and the proportion of patients with resting heart rate >87bpm.

### Explanatory analyses

The associations between clinical characteristics before surgery, morbidity, autonomic function and heart rate changes from preoperative values outcomes were investigated.

### Statistical analysis

Normally distributed continuous data are presented as mean (standard deviation) and compared using the paired t-test, repeated measures or one-way analysis of variance (ANOVA), as indicated in the text. Skewed data are presented as median (interquartile range) and compared using the Wilcoxon signed-rank test. Categorical data are presented as numerical values and compared using Chi-square tests. For explanatory analyses of morbidity after surgery, quartiles of absolute discharge heart rate were compared. Multivariable regression analysis explored the relationship between discharge heart rate and perioperative factors that may influence heart rate, including age, sex, body-mass index, preoperative heart rate, Revised Cardiac Risk Index, frequency-domain measures of heart rate variability and postoperative morbidity. Two-sided P values <0.05 were considered significant. All statistical analyses were performed with NCSS 11 (Kaysville, Utah USA).

### Sample size calculation

To detect a mean (SD) increase in resting heart rate at discharge of 10 (5) beats minute^-1^, compared to preoperative heart rate, at least 73 patients would be required (1-β=0.1; α=0.05).

## Results

### Patient characteristics

Two hundred and ten patients were enrolled between 5^th^ October 2016 to 4^th^ December 2018. Fifty-patients did not have discharge heart rate recorded, leaving 157 patients in this analysis (Figure 1). Mean length of hospital stay was 8 days (interquartile range 4-11 days) after surgery (Table 1). No patients died within 30 days of surgery.

**Figure 1.**
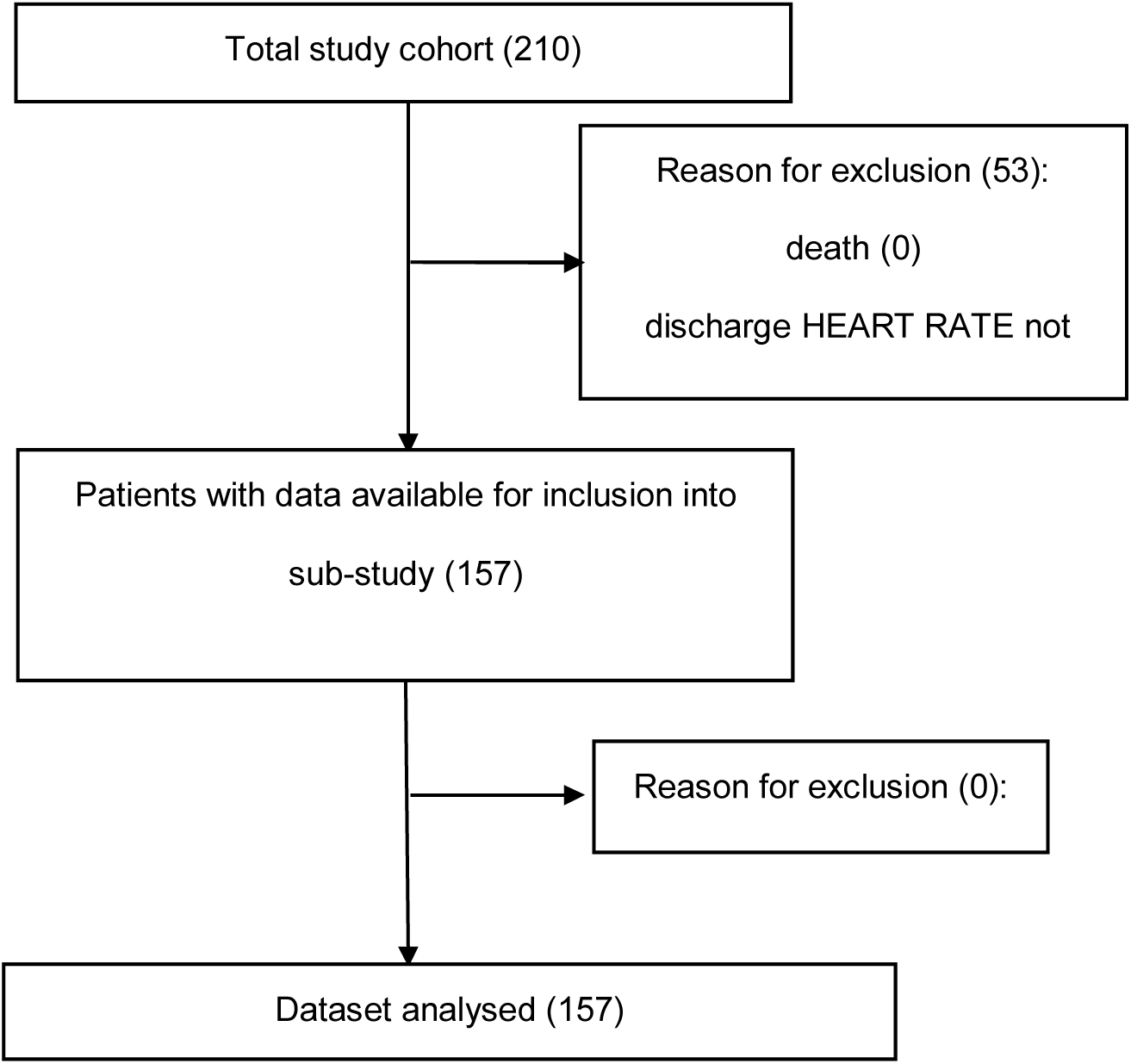
Study flow and analysis.

**Table 1:**
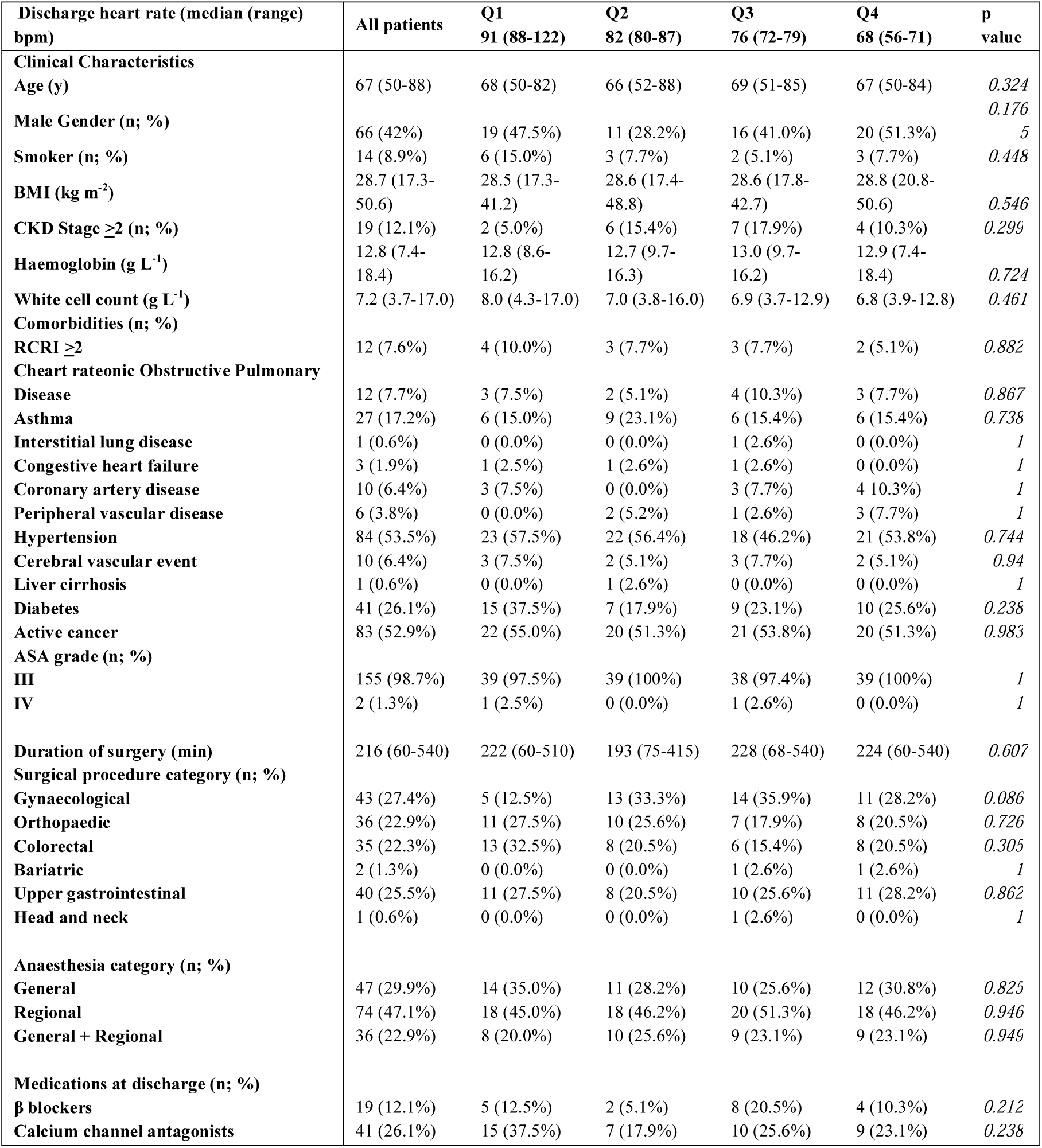
Baseline characteristics. Data is presented as mean with standard deviations (SD) for parametric data and as median (25th-75th interquartile range) for non-parametric data. Frequencies are presented with percentages (%). Age is rounded to the nearest year. CKD: Chronic kidney disease; stage is defined as per KDIGO recommendations (15). RCRI: Revised cardiac risk index score. ASA: American Association of Anesthesiologists. Other units as indicated. Statistical analysis using Student’s unpaired t-test for continuous data and the χ^2^ test for categorical data.

### Primary outcome: absolute discharge heart rate

Heart rate at discharge ranged from 56-122 beats minute^-1^, with mean heart rate 80 (SD 12) beats minute^-1^.

### Secondary outcomes: heart rate changes from preoperative values

At hospital discharge, mean heart rate was 5 beats minute^-1^ (95%CI:3-7) higher, compared to heart rate values recorded at pre-assessment clinic (P<0.001, by paired t-test). Heart rate changes from preoperative assessment values ranged from 28 beats minute^-1^ lower, to 39 beats minute^-1^ higher, at the time of discharge from hospital (Figure 2). Qualitatively similar results were seen when we compared preoperative heart rate on the day of surgery, recorded by Holter monitoring (mean heart rate difference: 11 beats minute^-1^ (95%CI:9-13); P<0.001).

**Figure 2.**
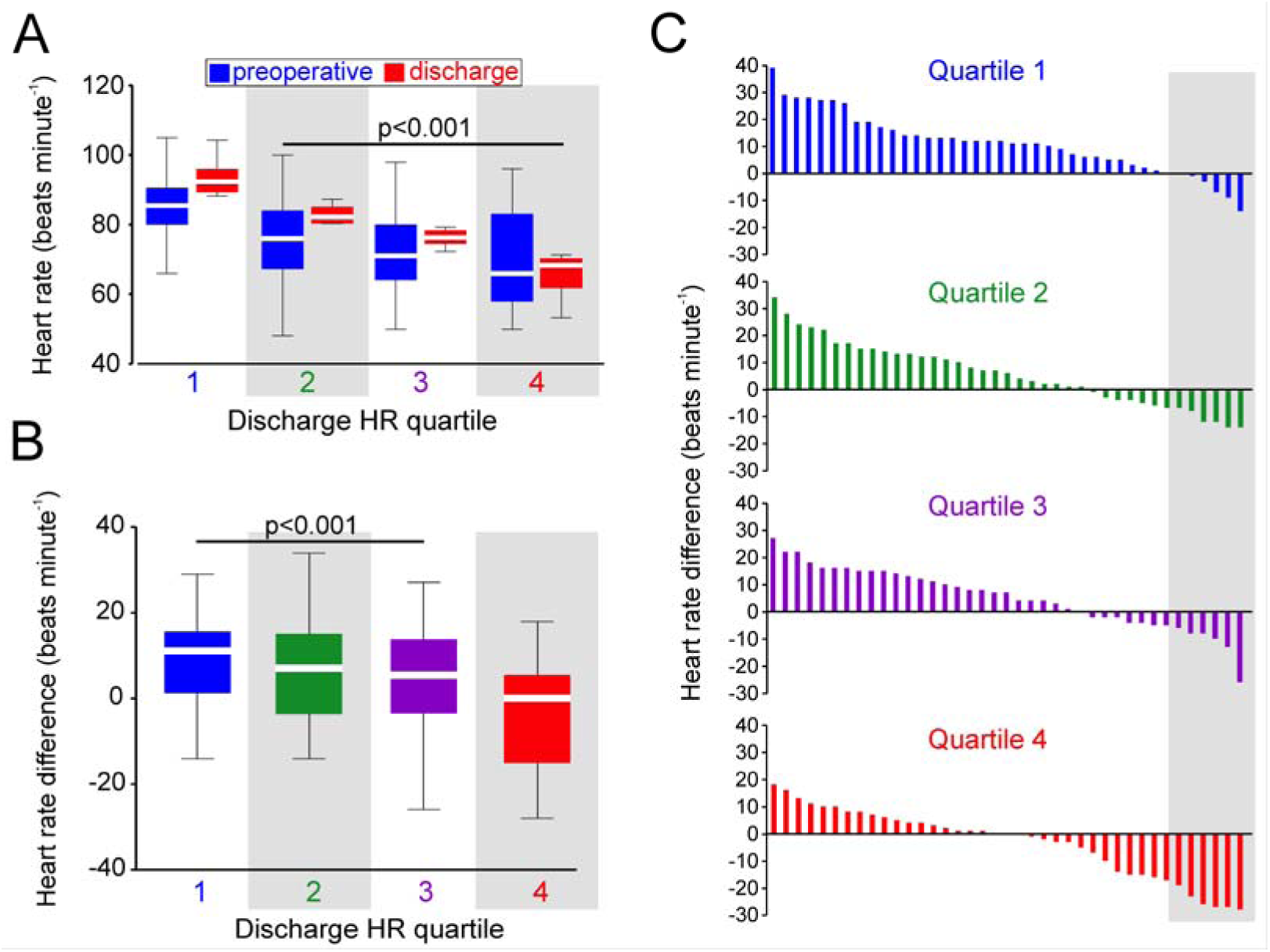
Perioperative changes in heart rate. A. Median preoperative and discharge heart rates, arranged by quartiles of discharge heart rate. P value derived by one-way ANOVA, comparing between quartiles. B. Median changes in heart rate at discharge from preoperative values, arranged by quartiles of discharge heart rate. P value derived by one-way ANOVA, comparing between quartiles. C. Waterfall plots showing change in heart rate from preoperative to hospital discharge values for each quartile of discharge heart rate.

Mean preoperative heart rate in patients in the upper quartile of discharge heart rate (85 (95%CI:82-88)) was 9 beats minute^-1^ higher (95%CI:2-16); P<0.001), compared to the other three quartiles (Figure 2A). Patients with a resting discharge heart rate >87bpm had a higher preoperative heart rate (84 beats minute^1^ (95%ci: 3-12); p=0.001), but also sustained a greater increase in heart rate after surgery (+11 beats minute^-1^ (95% ci:7-15); p<0.0001), compared to patients with a discharge heart rate ≤87bpm (Figure 2B).

### Discharge heart rate and morbidity within first 7 days of surgery

Patients in the upper quartile of discharge heart rate (>87bpm) were more likely to sustain pulmonary (OR:2.18 (95%CI:1.07-4.44); p=0.03), and infectious (OR:2.31 (95%CI:1.13-4.75); p=0.02) morbidity within seven days of surgery, compared to lower quartiles (Table 2). A similar incidence of myocardial injury (OR:1.17 (95%CI:0.52-2.65); p=0.68) and POMS-defined cardiovascular morbidity (OR:1.01 (95%CI:0.45-2.26); p=0.68) was observed between patients in the upper quartile of discharge heart rate (>87bpm), compared to lower quartiles. Patients in the upper quartile of discharge heart rate (>87bpm) stayed in hospital for longer after surgery (Supplementary data).

**Table 2.** Morbidity within first 7 days of surgery. Morbidity defined by Postoperative Morbidity Survey. Number of patients (%) shown for each quartile of discharge heart rate. Absolute heart rate values shown as (median (range)).

|  | Quartile 1 | Quartile 2 | Quartile 3 | Quartile 4 | Odds ratio<br>(95%CI) | P<br>value |
| --- | --- | --- | --- | --- | --- | --- |
| **Discharge heart rate** | 91 (88-122) | 82 (80-87) | 76 (72-79) | 68 (56-71) | --- | --- |
| **Pulmonary** | 26 (59%) | 14 (39%) | 20 (50%) | 11 (30%) | 2.18 (1.07-4.44) | 0.03 |
| **Infectious** | 21 (48%) | 8 (22%) | 14 (35%) | 10 (27%) | 2.31 (1.13-4.75) | 0.02 |
| **Renal** | 23 (52%) | 13 (36%) | 20 (50%) | 18 (49%) | 1.33 (0.66-2.68) | 0.42 |
| **Gastrointestinal** | 21 (48%) | 12 (33%) | 16 (40%) | 15 (41%) | 1.49 (0.74-3.00) | 0.27 |
| **Cardiovascular** | 11 (25%) | 8 (22%) | 15 (38%) | 5 (14%) | 1.01 (0.45-2.26) | 0.98 |
| **Neurological** | 5 (11%) | 3 (8%) | 2 (5%) | 2 (5%) | 1.94 (0.58-6.48) | 0.28 |
| **Haematological** | 4 (9%) | 1 (3%) | 1 (3%) | 2 (5%) | 2.73 (0.65-11.4) | 0.17 |
| **Wound** | 1 (2%) | 0 (0%) | 0 (0%) | 0 (0%) | 7.83 (0.31-195) | 0.21 |
| **Pain** | 20 (45%) | 13 (36%) | 20 (50%) | 12 (32%) | 1.26 (0.62-2.54) | 0.52 |

### Cardiac autonomic function

Heart rate was similar between patients who sustained, or avoided, early pulmonary and/or infectious morbidity (Figure 3A-C). Time-domain HRV analysis revealed lower SDNN in patients who sustained pulmonary and infectious morbidity within 5 days of undergoing surgery (Figure 3D-F). Analysis of frequency-domain measures revealed that both lnLF (p=0.02) and lnHF (p=0.03) were reduced in patients who sustained pulmonary and infectious POMS-defined complications after surgery (Figure 3G-J).

**Figure 3.**
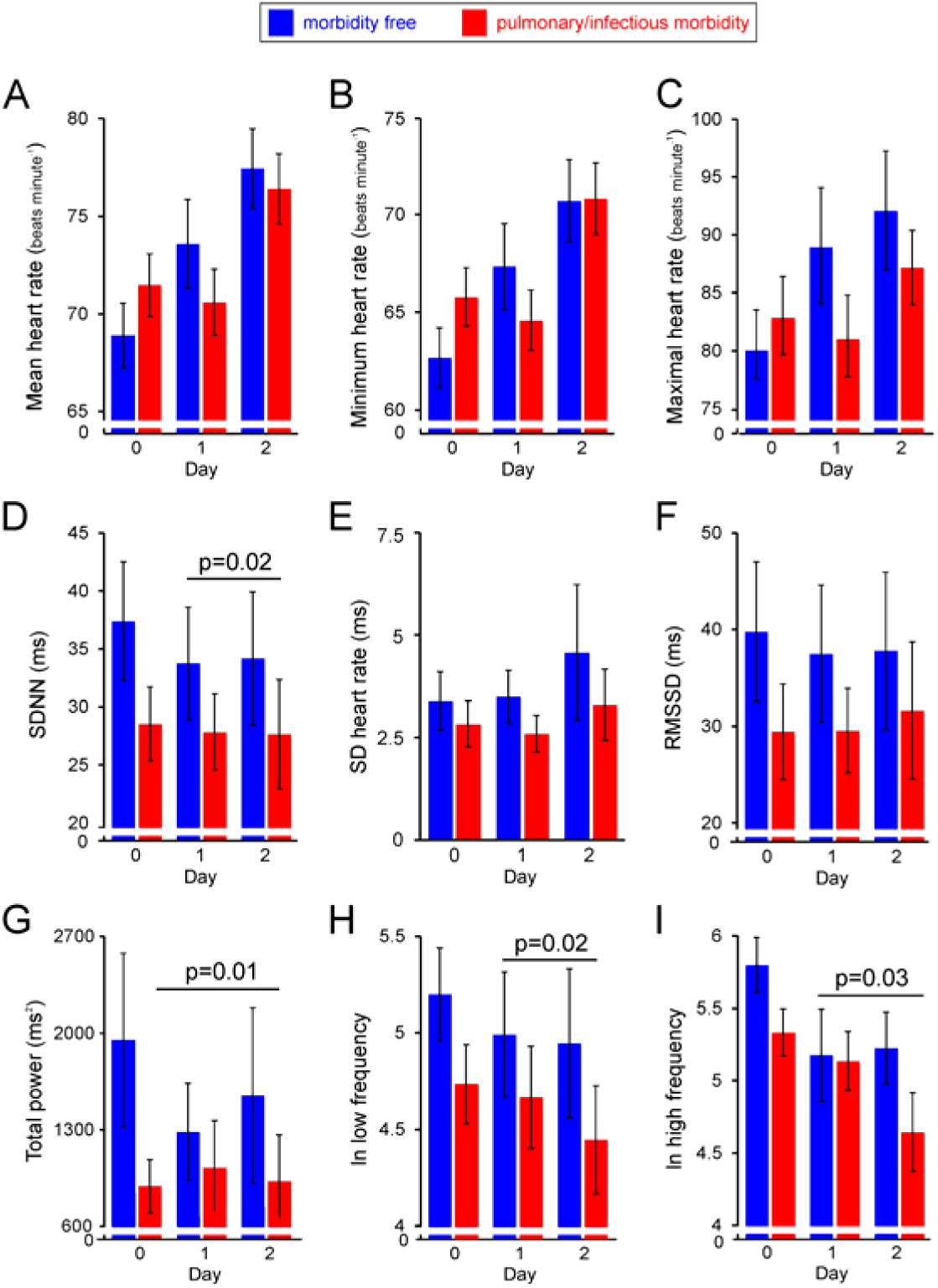
Time/frequency domain measures of heart rate variability associated with morbidity after surgery. A-F. Serial time-domain HRV measures within first 48h of surgery, compared between patients who acquired pulmonary and/or infectious morbidity. A. mean heart rate. B. minimum heart rate. C. maximal heart rate. D. SDNN. E. SD. F. RMSSD. G-I. Frequency-domain HRV measures within first 48h of surgery, compared between patients who acquired pulmonary and/or infectious morbidity. G. Total power. H. lnLF. I. lnHF. P values denote post-hoc Tukey testing, following repeated-measures ANOVA (day x morbidity)

### Multivariable analysis of discharge heart rate

Older age (OR:1.07 (95%CI:1.02-1.13); p=0.01) and lower preoperative heart rate (OR:0.89 (95%CI:0.86-0.94); p<0.001) were independently associated with the highest heart rate quartile at discharge, accounting for resumption of beta-blockade before leaving hospital, morbidity after surgery, Revised Cardiac Risk Index and perioperative changes in cardiac autonomic control (Supplementary data)

## Discussion

We found that resting heart rate within 24h of discharge from hospital following major non-cardiac surgery was frequently elevated, compared to resting preoperative values. The most elevated discharge heart rate was associated with higher preoperative heart rate and cardiac vagal tone, as reflected by low frequency-domain fast fourier transform analysis. There was no association between use of beta/calcium channel blocking agents at discharge. Given the well-established and mechanistically robust association between heart rate and development of cardiovascular (and extra-cardiovascular) pathology, discharge heart rate appears to be a plausible modifiable risk factor for delayed recovery after noncardiac surgery.

Discharge heart rate is an independent predictor of morbidity and mortality in different populations treated for acute myocardial infarction (3,4,5), and heart failure (2,6). Patients with myocardial infarction and left ventricular failure have demonstrated an independent association between resting heart rate and risk of overall mortality during 10-year follow-up (16). This notion is supported by a study of patients treated for ST-elevation myocardial infarction patients by primary PCI, in which the association between elevated discharge heart rate and increased 4-year mortality was independent of myocardial infarction size and/or the development of heart failure (4). Elevated heart rate increases myocardial oxygen consumption (17), and reduces diastolic filling time (and hence coronary perfusion) (18). Raised heart rate also augments atherosclerosis severity and progression in experimental models, with patients at higher risk of coronary plaque rupture (19). heart rate reduction has attenuated atherosclerosis progression in experimental models, suggesting that therapeutic heart rate reduction may improve endothelial function and confer cardioprotective effects (20). Other proposed pathophysiological consequences of elevated heart rate include reduced development of collaterals vessels and increased arterial rigidity (20). These processes may subsequently lead to left ventricular dysfunction and a higher incidence of arrhythmias (21,22), contributing to non-cardiac and cardiac morbidity and mortality. The randomized placebo-controlled Systolic Heart failure treatment with the If inhibitor ivabradine (SHIFT) trial investigated the association between heart rate and cardiovascular events in heart failure, by treating a chronic heart failure patient population with the selective heart-rate-lowering agent ivabradine versus placebo. Selective heart rate reduction with ivabradine lowered the risk of adverse cardiovascular outcomes.(23) This indicates that heart rate greatly contributes to the progression of heart failure, suggesting that heart rate reduction should be a key therapeutic target (24).

Major surgery may result in elevated heart rate even in the absence of key stimuli for chemoreceptor-driven cardiovascular responses, evoked by postoperative hypoxia, hypercapnia and acidosis (25), which quickly alter heart rate variability (26). Our study shows that elevated discharge heart rate after elective non-cardiac surgery is associated with pulmonary and infectious morbidity after surgery. This finding is consistent with a modulatory role of inflammation on autonomic control. Activation of toll-like receptors (TLRs) in the peripheral carotid chemoreceptors by inflammatory danger-associated molecular patterns (DAMPs) stimulates chemoreceptor-driven autonomic responses including tachycardia, even in the absence of hypoxia. (27)

Our exploratory analysis identified resting heart rate before surgery to be strongly associated with higher discharge heart rate. Cardiac vagal tone is the main determinant of resting heart rate. (28) From VISION studies, resting heart rate >87 beats minute^-1^ before surgery is independently associated with myocardial injury.(8) A substantial number of patients either entered surgery with heart rate>87 beats minute^-1^ or breached this prognostic theart rateeshold by the time of discharge. Moreover, the mean heart rate value within the highest heart rate quartile was ∼87 beats minute^-1^, suggesting that patients discharged with an elevated heart rate following major elective surgery may be at increased risk of subsequent morbidity, including cardiovascular and thrombotic complications that occur for at least 12 months after noncardiac surgery.(29) Elevated discharge heart rate may therefore be a modifiable risk factor for enhancing recovery after noncardiac surgery.

The main strength of this prospective study was the capture of both resting preoperative and discharge heart rate values, although we acknowledge that time-weighted measures are more likely to accurately reflect heart rate changes after surgery. Nevertheless, patients and clinicians were blinded to perioperative data and outcomes, which included an established tool to measure perioperative morbidity. Although heart rate was measured objectively using Holter monitors, recordings were made within 72h of surgery and may therefore not reflect changes in heart rate thereafter. We could not standardise how heart rate was measured by clinical staff, although patients had heart rate recordings in the sitting position. We cannot also control for diurnal variation in heart rate (30), although this may be abolished in any event by systemic inflammation synonymous with surgery.

## Conclusion

On discharge from hospital after major elective non-cardiac surgery, heart rate is elevated compared to resting preoperative values. Higher discharge heart rate is associated with prognostically relevant autonomic dysfunction before, and inflammatory morbidity after, surgery. Targeting elevated heart rate at discharge from hospital, which is a modifiable risk factor, may accelerate recovery after elective non-cardiac surgery.

## Data Availability

all enquiries to corresponding author

## Author contributions

Conceived and designed the study: G.L.A. Performed the experiments: S.M.M., A.R., G.M. Designed the analysis, analysed the data, and wrote the first draft of the manuscript: A.B.U.P., G.L.A. Contributed to the writing of the manuscript: A.B.U.P., S.M.M., T.E.F.A., G.L.A. Agree with manuscript results and conclusions: A.B.U.P., S.M.M., T.E.F.A., A.R., G.M., G.L.A.

## SUPPLEMENTARY MATERIALS

### STROBE compliance

We complied with The Strengthening and Reporting of Observational Studies in Epidemiology Statement (11), as illustrated below. *Give information separately for exposed and unexposed groups.

|  | Item | Recommendation | Page |
| --- | --- | --- | --- |
| **Title and abstract** | 1 | (a) Indicate the study's design with a commonly used term in the title or the abstract<br>(b) Provide in the abstract an informative and balanced summary of what was done and what was found | 2 |
| **Introduction** |  |  |  |
| Background/rationale | 2 | Explain the scientific background and rationale for the investigation being reported | 3 |
| Objectives | 3 | State specific objectives, including any prespecified hypotheses | 3 |
| **Methods** |  |  |  |
| Study design | 4 | Present key elements of study design early in the paper | 4 |
| Setting | 5 | Describe the setting, locations, and relevant dates, including periods of recruitment, exposure, follow-up, and data collection | 4 |
| Participants | 6 | (a) Give the eligibility criteria, and the sources and methods of selection of participants. Describe methods of follow-up<br>(b) For matched studies, give matching criteria and number of exposed and unexposed | 4 |
| Variables | 7 | Clearly define all outcomes, exposures, predictors, potential confounders, and effect modifiers. Give diagnostic criteria, if applicable | 4 |
| Data sources/ measurement | 8* | For each variable of interest, give sources of data and details of methods of assessment (measurement). Describe comparability of assessment methods if there is more than one group | 5-8 |
| Bias | 9 | Describe any efforts to address potential sources of bias | 7 |
| Study size | 10 | Explain how the study size was arrived at | 8 |
| Quantitative variables | 11 | Explain how quantitative variables were handled in the analyses. If applicable, describe which groupings were chosen and why | 5-8 |
| Statistical methods | 12 | (a) Describe all statistical methods, including those used to control for confounding<br>(b) Describe any methods used to examine subgroups and interactions<br>(c) Explain how missing data were addressed<br>(d) If applicable, explain how loss to follow-up was addressed<br>(e) Describe any sensitivity analyses | 7-8 |
| **Results** |  |  |  |
| Participants | 13* | (a) Report numbers of individuals at each stage of study—eg numbers potentially eligible, examined for eligibility, confirmed eligible, included in the study, completing follow-up, and analysed<br>(b) Give reasons for non-participation at each stage<br>(c) Consider use of a flow diagram | 9 |
| Descriptive data | 14* | (a) Give characteristics of study participants (eg demographic, clinical, social) and information on exposures and potential confounders<br>(b) Indicate number of participants with missing data for each variable of interest<br>(c) Summarise follow-up time (eg, average and total amount) | 9 |
| Outcome data | 15* | Report numbers of outcome events or summary measures over time | 9 |
| Main results | 16 | (a) Give unadjusted estimates and, if applicable, confounder-adjusted estimates and their precision (eg, 95% confidence interval). Make clear which confounders were adjusted for and why they were included<br>(b) Report category boundaries when continuous variables were categorized<br>(c) If relevant, consider translating estimates of relative risk into absolute risk for a meaningful time period | 10 |
| Other analyses | 17 | Report other analyses done—eg analyses of subgroups and interactions, and sensitivity analyses | 10 |
| **Discussion** |  |  |  |
| Key results | 18 | Summarise key results with reference to study objectives | 12 |
| Limitations | 19 | Discuss limitations of the study, taking into account sources of potential bias or imprecision. Discuss both direction and magnitude of any potential bias | 14 |
| Interpretation | 20 | Give a cautious overall interpretation of results considering objectives, limitations, multiplicity of analyses, results from similar studies, and other relevant evidence | 12 |
| Generalisability | 21 | Discuss the generalisability (external validity) of the study results | 12 |
| **Other information** |  |  |  |
| Funding | 22 | Give the source of funding and the role of the funders for the present study and, if applicable, for the original study on which the present article is based | 15 |

### Holter monitoring

#### Processing Electrocardiogram recording

Electrocardiogram recordings were made using the Lifecard CF digital Holter monitoring (Spacelabs Healthcare, Hertford UK) with a sampling rate of 1024Hz. The recordings were uploaded onto Sentinel V9 Cardiology information software system (Spacelabs Healthcare, Hertford, UK) and *.ecg files were then exported and loaded onto Kubios 3.1 software. All electrocardiogram recordings were visually inspected for quality of recording. 5 min (300s) epochs were selected for heart rate variability analysis after 2.5 min resting time to minimise movement artefact and to maximise a stationary time series.

#### R-R time series

5 min R-R interval time series were generated using the Kubios software in-built QRS detection algorithm. After visual inspection, missed beats were manually added and/or incorrect beats deleted. Patients whose ECG recordings could not generate a R-R interval times series due to no electrical recordings were excluded from further analysis.

#### Artefact detection and correction

After manual correction, ECG recordings with artefact or ectopic beats were processed using automatic artefact detection and correction where detected ectopic beats, or too long or short beats are corrected by replacing the R-R interval times with interpolated values. To account for slow non nonstationarities in the R-R interval detrending using smoothing priors λ = 500 was employed. The mean percentage of beats corrected per sample for this study was 1.8%. Time series where the % artefact correction was >5% were excluded from the analysis (unsuitable quality).

#### Heart rate variability- time domain analysis

Time domain and frequency domain analyses were performed on 300s R-R time series using Kubios 3.1 software after manual inspection +/- automatic correction. Statistical analyses derived directly from the 300s R-R interval time series used in this study were:Mean heart rate, Standard Deviation of the RR interval (SDNN) and Square root of the, mean squared differences between successive RR intervals (RMSSD). SDNN has previously been shown to be a predictor of mortality in chronic heart failure patients and post myocardial infarction. RMSSD is a measure of short term variation and correlate with parasympathetic modulation.

#### Heart rate variability - frequency domain analyses

Cubic spline interpolating at the sampling rate 4Hz was used to convert the R-R time series into an equidistant sampled series prior to spectrum estimation. Fast Fourier time spectrum was calculated using Welch’s Perdiodogram method (window width 128s with 50% overlap). The frequency bandwidths used were low frequency (LF) 0.04-0.15Hz and high frequency 0.15-0.4Hz. The frequency measures reflect autonomic modulation of heart rate variability. Power spectral density of HF is strongly associated with parasympathetic modulation of the heart. LF power spectral density has been shown to better correspond with baroreceptor function rather than sympathetic function. The absolute power (ms^2^) values were natural log transformed for statistical analysis.

**Suppl Table 1:** Postoperative morbidity survey (POMS) used to assess post-surgery morbidity in the study (13).

| Morbidity type | Criteria |
| --- | --- |
| Pulmonary | *De novo* requirement for supplemental oxygen or other respiratory support (e.g., CPAP or IPPV) |
| Infectious | Currently on antibiotics or temperature >38°C in the last 24 heart rates |
| Renal | Presence of oliguria (<500 mL/day), increased serum creatinine (>30% from baseline value), or urinary catheter in place for a non-surgical reasons |
| Gastrointestinal | Unable to tolerate an enteral diet (either by mouth or feeding tube) for any reason, including nausea, vomiting and abdominal distension |
| Cardiovascular | Diagnostic test or therapy in last 24 heart rates for any of the following reasons:<br>*de novo* myocardial infarction or ischemia, hypotension (requiring drug therapy or fluid >200 mL/heart rate), atrial or ventricular arrhythmia or pulmonary edema |
| Neurological | Presence of a *de novo* focal deficit, coma or confusion/delirium |
| Wound complications | Wound dehiscence requiring surgical exploration or drainage or pus from the wound |
| Haematological | Requirement for any of the following within last 24 heart rates: blood, platelets, fresh frozen plasma or cryoprecipitate |
| Pain | Surgical wound pain significant enough to require parenteral opiates or regional anaesthesia. |

